# Changes in population immunity against infection and severe disease from SARS-CoV-2 Omicron variants in the United States between December 2021 and November 2022

**DOI:** 10.1101/2022.11.19.22282525

**Authors:** Fayette Klaassen, Melanie H. Chitwood, Ted Cohen, Virginia E. Pitzer, Marcus Russi, Nicole A. Swartwood, Joshua A. Salomon, Nicolas A. Menzies

## Abstract

**Importance:** While a substantial fraction of the US population was infected with SARS-CoV-2 during December 2021 – February 2022, the subsequent evolution of population immunity against SARS-CoV-2 Omicron variants reflects the competing influences of waning protection over time and acquisition or restoration of immunity through additional infections and vaccinations.

**Objective:** To estimate changes in population immunity against infection and severe disease due to circulating SARS-CoV-2 Omicron variants in the United States from December 2021 to November 2022, and to quantify the protection against a potential 2022-2023 winter SARS-CoV-2 wave.

**Design, setting, participants:** Bayesian evidence synthesis of reported COVID-19 data (diagnoses, hospitalizations), vaccinations, and waning patterns for vaccine- and infection-acquired immunity, using a mathematical model of COVID-19 natural history.

**Main Outcomes and Measures:** Population immunity against infection and severe disease from SARS-CoV-2 Omicron variants in the United States, by location (national, state, county) and week.

**Results:** By November 9, 2022, 94% (95% CrI, 79%–99%) of the US population were estimated to have been infected by SARS-CoV-2 at least once. Combined with vaccination, 97% (95%–99%) were estimated to have some prior immunological exposure to SARS-CoV-2. Between December 1, 2021 and November 9, 2022, protection against a new Omicron infection rose from 22% (21%–23%) to 63% (51%–75%) nationally, and protection against an Omicron infection leading to severe disease increased from 61% (59%–64%) to 89% (83%–92%). Increasing first booster uptake to 55% in all states (current US coverage: 34%) and second booster uptake to 22% (current US coverage: 11%) would increase protection against infection by 4.5 percentage points (2.4–7.2) and protection against severe disease by 1.1 percentage points (1.0–1.5).

**Conclusions and Relevance:** Effective protection against SARS-CoV-2 infection and severe disease in November 2022 was substantially higher than in December 2021. Despite this high level of protection, a more transmissible or immune evading (sub)variant, changes in behavior, or ongoing waning of immunity could lead to a new SARS-CoV-2 wave.

**Key points:** *Question:* How did population immunity against SARS-CoV-2 infection and subsequent severe disease change between December 2021, and November 2022?

*Findings:* On November 9, 2022, the protection against a SARS-CoV-2 infection with the Omicron variant was estimated to be 63% (51%–75%) in the US, and the protection against severe disease was 89% (83%–92%).

*Meaning:* As most of the newly acquired immunity has been accumulated in the December 2021-February 2022 Omicron wave, risk of reinfection and subsequent severe disease remains present at the beginning of the 2022-2023 winter, despite high levels of protection.

## Introduction

Three years into the epidemic, SARS-CoV-2 is still circulating worldwide. Over the course of 2022, the Omicron subvariants B.1.1.529 and BA.5 emerged and became dominant in the United States^1^, and several new subvariants have subsequently emerged with potential to outcompete the current dominant variants. During the first Omicron wave (December 2021 – February 2022) over 25 million COVID-19 diagnoses were reported in the US, equivalent to 8% of the population. Many more infections were likely undiagnosed and, due to waning immunity and increased immune evasion, reinfections are occurring more frequently^2^, even amongst those vaccinated^3^.

COVID-19 cases are expected to rise again in the United States towards the end of 2022, co-occurring with seasonal influenza and other respiratory viruses. Over the course of 2022, mask mandates and restrictions on social gatherings have been lifted, reducing barriers to the transmission of respiratory pathogens. In the absence of non-pharmaceutical interventions, partial immunity afforded by previous infection and vaccination is the main protection against future infections. By November 9, 2022, 79% of the US population had initiated a vaccination series, 34% had received a first booster and 11% had received a second booster^1^. Levels of vaccination coverage have been heterogeneous geographically and temporally: over three-quarters of the vaccinations in 2022 were administered in January, and the fraction of the population who had received any booster by November 9, 2022 ranged from 20% in North Carolina to 55% in Vermont.

Ongoing (re)infections and vaccinations boost population immunity against future infections and subsequent severe disease. These processes are offset by waning of immunity as well as immune evasion of newer variants. Estimates of changes in population immunity need to account for these competing mechanisms. Modeling this process is complicated by the emergence of variants with different inherent transmissibility and immune evasion properties. The levels of protection conferred by different exposure types and the rate at which that protection wanes remains uncertain. With the emergence of the Omicron variant, vaccine protection was strongly reduced as compared to earlier variants^4-6^. While receiving a booster dose increases the protection against infection^7^, with a stronger effect for the bivalent booster^8^, the low uptake to date limits this protection in the population. Previous infection also offers less protection against reinfection with the Omicron variant compared to earlier variants^9^, although Omicron infections offer more protection than pre-Omicron infections^10^. Evidence suggests that protection afforded by a prior infection is higher than that acquired by vaccination alone, and that hybrid immunity offers the most durable protection^11^. As a consequence of these different factors, the US population at the end of 2022 will reflect a range of prior immunological exposures to SARS-CoV-2, with variable protection against future infections.

In this study we combined estimates of infection and vaccination trends with recent evidence on temporal patterns in immunity, to estimate the level of population immunity in the US at national, state, and county levels, for each week between 1 December 2021 and 9 November 2022.

## Methods

### Data

We extracted CDC data for state- and county-level vaccination and booster coverage between December 2, 2021 and November 9, 2022^1^. We consider three vaccination states. First, we used the reported number of individuals that initiated a primary vaccination series as an indicator of being vaccinated. Second, we used the reported number of individuals that received an additional dose after having completed the primary series as an indicator of being boosted. Thirdly, we used the reported number of individuals that received a second booster shot, after having received a primary vaccination series and a first booster dose. Because this data was unavailable at the county level, we assumed the second booster coverage was equal to the encompassing state.

We used a statistical model that renders weekly time-series of SARS-CoV-2 first infections and reinfections, accounting for both under-ascertainment and time lags of reporting and disease progression^12^. Inputs to this model are reported case^13,14^ and hospitalization data^15^. The published model has been updated to better represent disease dynamics under the Omicron variants of SARS-CoV-2 in three ways. First, the updated model allows for waning of infection-induced immunity and includes a reduced probability of progressing to more severe disease states for those reinfected^1,16^. Second, the model was fitted to hospitalization reports^15^, rather than deaths, due to the reduced infection fatality ratio and subsequent sparser deaths due to SARS-CoV-2 over 2022. To facilitate this change, the updated model renders weekly rather than daily estimates. Third, the infection fatality ratio has been updated to reflect the decreased mortality risk under the Omicron variants (additional eMethods provided in the Supplement). The model used for this analysis was initialized with immunity estimates as of December 2, 2021, as reported by Klaassen et al. (2022)^17^.

### Estimates of immunological exposure

For each location and each week, we computed the percentage of the population *immunologically exposed*, defined having received at least one dose of a COVID-19 vaccine, at least one prior SARS-CoV-2 infection, or both, using the cumulative reports of first infections and first vaccine doses. Following Klaassen et al. (2022),^17^ we operationalized the probability of co-occurrence of infection and vaccination as odds ratios for vaccination among individuals with vs. without prior infection, based on data collected in the CDC Household Pulse Surveys^18^ and validated against independent survey estimates^19^.

### Effective protection by exposure state

We defined exposure states classifying the population into mutually exclusive categories for different combinations of prior infection and vaccination (eTable 1). Based on the estimated immunological exposure and the waning curves, we estimated how individuals transition between these exposure states over time. At each timepoint, we determined the number newly entering the joint vaccinated and infected state through either infection or vaccination, and correspondingly calculated the fraction of the new infections and vaccinations occurring in the immune-naïve population. Next, we distributed the new reinfections over those previously infected. Finally, the booster and second booster doses were assigned proportionally over the vaccinated and first booster exposure states. We assumed that at each time point only one event could take place (e.g., it was not possible to be infected and receive a booster in the same week). Infections and reinfections were assigned across the exposures states proportional to susceptibility to (re)infection, while vaccinations and boosters were assigned proportional to exposure. We used the population distribution across these exposure states, combined with the waning rates associated with each state, to estimate the level of population immunity against infection and severe disease from an Omicron variant for each week and modelled location, between December 2, 2021 and November 9, 2022. At each time point, we adjusted for mortality and birth rates. Specifically, we distributed the deaths proportionally over all exposure states, and assigned the births to the naïve exposure state. We used birth and mortality reports for each US state and each month in 2021 to compute average weekly rates^20^. We assumed the birth and mortality rates for counties were equal to that of their encompassing state.

### Initial levels of protection and subsequent waning

A recent systematic review and meta-analysis synthesized evidence on the waning of protection against infection and severe disease from exposure to the Omicron variants^9,10,21^. We used this evidence to specify waning curves for different combinations of prior infection and vaccination. We assumed that immunity is lost at constant rate, such that each waning curve is parameterized as an initial level of immunity and an exponential rate of decline. The initial level of immunity ranges from 30% for individuals who had only received their primary series to 85% protection for individuals who have received a booster dose and been infected with the Omicron variant (eTable 2 in the Supplement). Based on the evidence reported in the meta-analysis, we assumed that immunity against infection declined at the same rate for all non-hybrid exposures ((re)infection only, vaccination/booster only), and at a lower rate for hybrid exposures (both prior infection and vaccination). For protection against severe disease, we assumed immunity declined at 1% per month and stayed constant for hybrid exposures. We set a lower bound for the protection against infection at 0.1 and for the protection against severe disease at 0.5, assuming some residual level of protection for individuals with any historical exposure compared to unexposed individuals. The waning curves derived from these assumptions are shown in Figure 1, along with the empirical estimates used to parameterize these curves.

**Figure 1:**
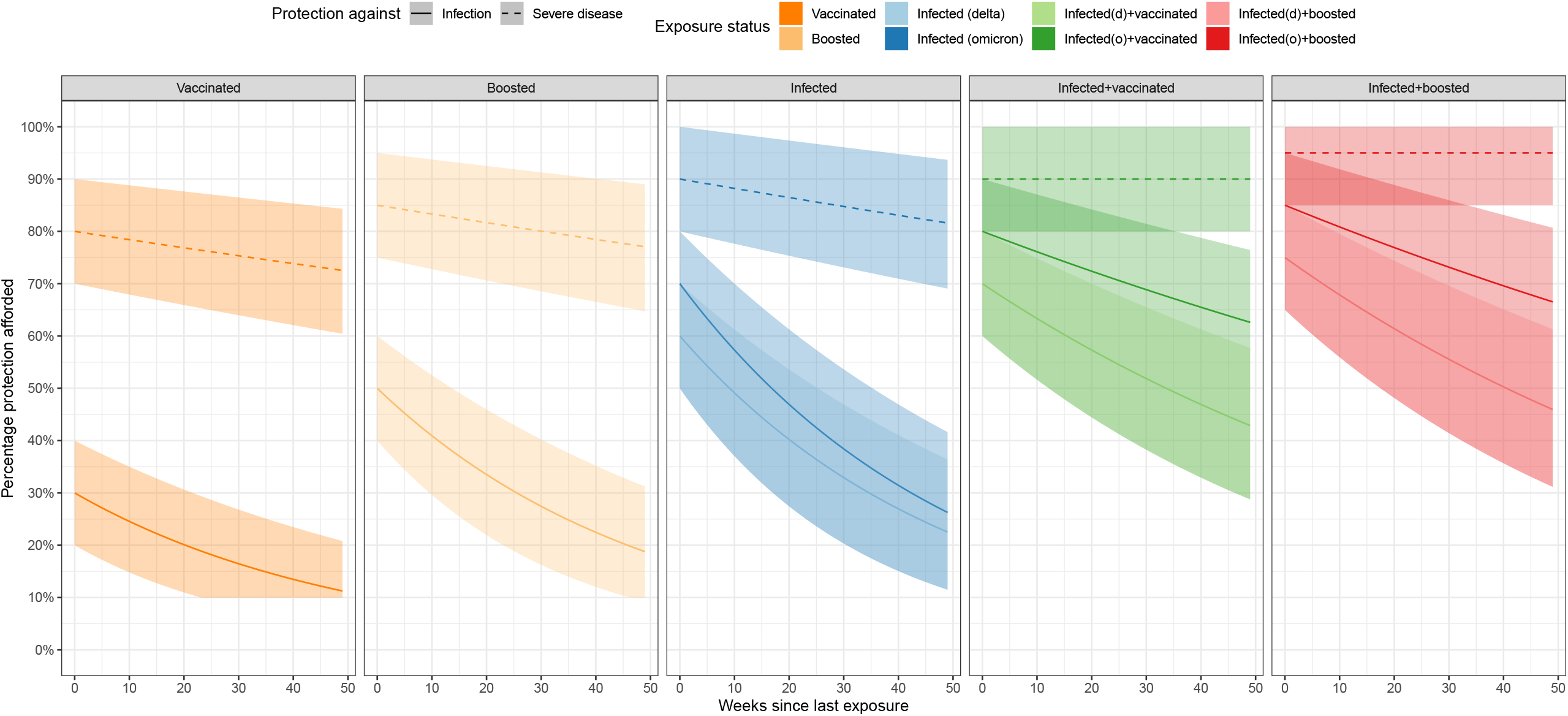
Assumed initial immunity and waning trends for different combinations of prior infection and vaccination

We compared the results of these assumptions to alternative scenarios with optimistic and pessimistic assumptions for initial immunity and waning rates. In the optimistic scenario, initial immunity estimates were assumed to be 10 percentage points higher than the base-case, and the rate of decline was 50% less. In the pessimistic scenario, initial immunity was assumed to be 10 percentage points lower than the base-case, and the rate of decline was 50% more. In this pessimistic scenario, we also assumed there was no additional benefit of having an Omicron infection in terms of the resulting immunity.

### Impact of higher booster uptake

We examined a final scenario estimating the potential impact of greater booster uptake. We operationalized this as an immediate increase in the population fraction that had received a COVID-19 booster and second booster (distributed proportionally across all exposure states eligible for each), from observed levels on November 9, 2022 to 55% coverage of first booster and 20% uptake of the second booster, which represent the highest values for these two outcomes across US states (both values taken from Vermont).

### Model implementation

We executed the analysis in R^22^ and the rstan package^23^ (https://github.com/covidestim/covidestim/tree/severe-protection). For state-level results, we report uncertainty using equal-tailed 95% credible intervals (95%CrI). We calculated national estimates and conservative uncertainty intervals by summing state-level estimates and upper and lower bounds of state-level intervals. County-level estimates were produced using an optimization routine^6^ that produces point estimates without uncertainty intervals. For 91 counties we could not compute estimates of immunity due to missing data.

## Results

### Immunological exposure

By November 9, 2022, 94% (95% CrI, 79%–99%) of the US population were estimated to have been infected by SARS-CoV-2 at least once, and 65% (35%–88%) were estimated to have experienced multiple infections over the course of the pandemic. Between December 2, 2021 and November 9, 2022, 116 (67–135) million first infections were estimated to have occurred, and 209 (118–292) million reinfections (corresponding to approximately 32% and 65% of the US population, respectively; on average, every person got infected 0.97 times). Combined with vaccination, 97% (95%–99%) were estimated to have some prior immunological exposure to SARS-CoV-2 by November 9, 2022. Figure 2 shows how the US population was assumed to transition through exposure states between December 2, 2021 and November 9, 2022. eFigures 1 and 2 show the immunological exposure states over time between December 2, 2021 and November 9, 2022 at national and state level, respectively.

**Figure 2:**
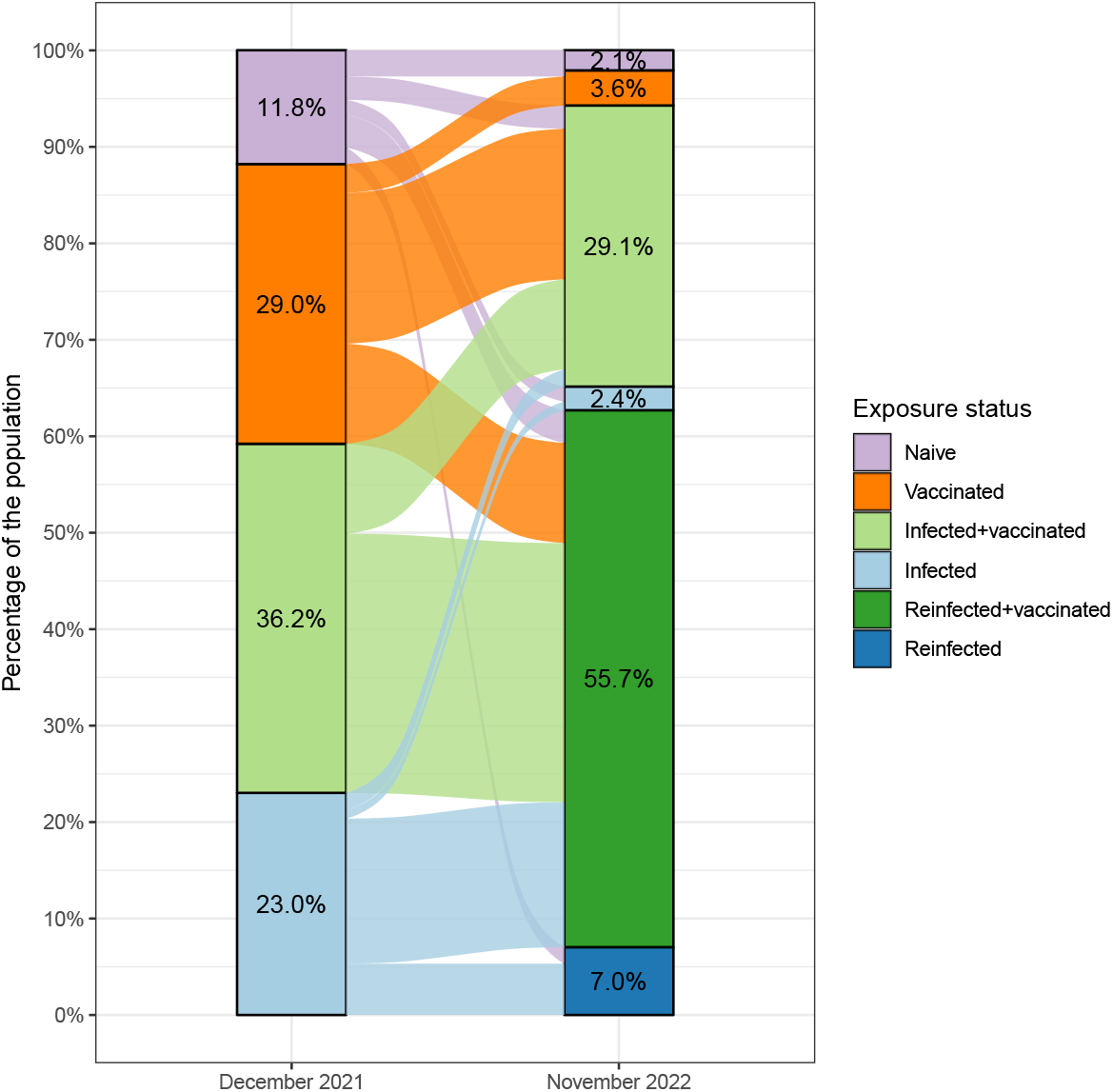
Flow of the US population between COVID-19 starting exposure states on December 1, 2021 to ending exposure state on November 9, 2022. Footnote: Individuals may make multiple transitions over this time period.

### Population immunity against infection

Between December 2, 2021 and November 9, 2022, population protection against an Omicron infection rose from 22% (21%–23%) to 63% (51%–75%) nationally (Figure 3AB, eFigure3, eFigure5). In contrast, population immunity against infection by the then circulating pre-Omicron variants in the fall of 2021 (November 9) was estimated to be 51% (47%–56%)^17^. Most of the additional population immunity accrued over the study period was acquired during the initial Omicron surge, with protection against infection estimated as 57% (47%–69%) on March 3, 2022.

**Figure 3:**
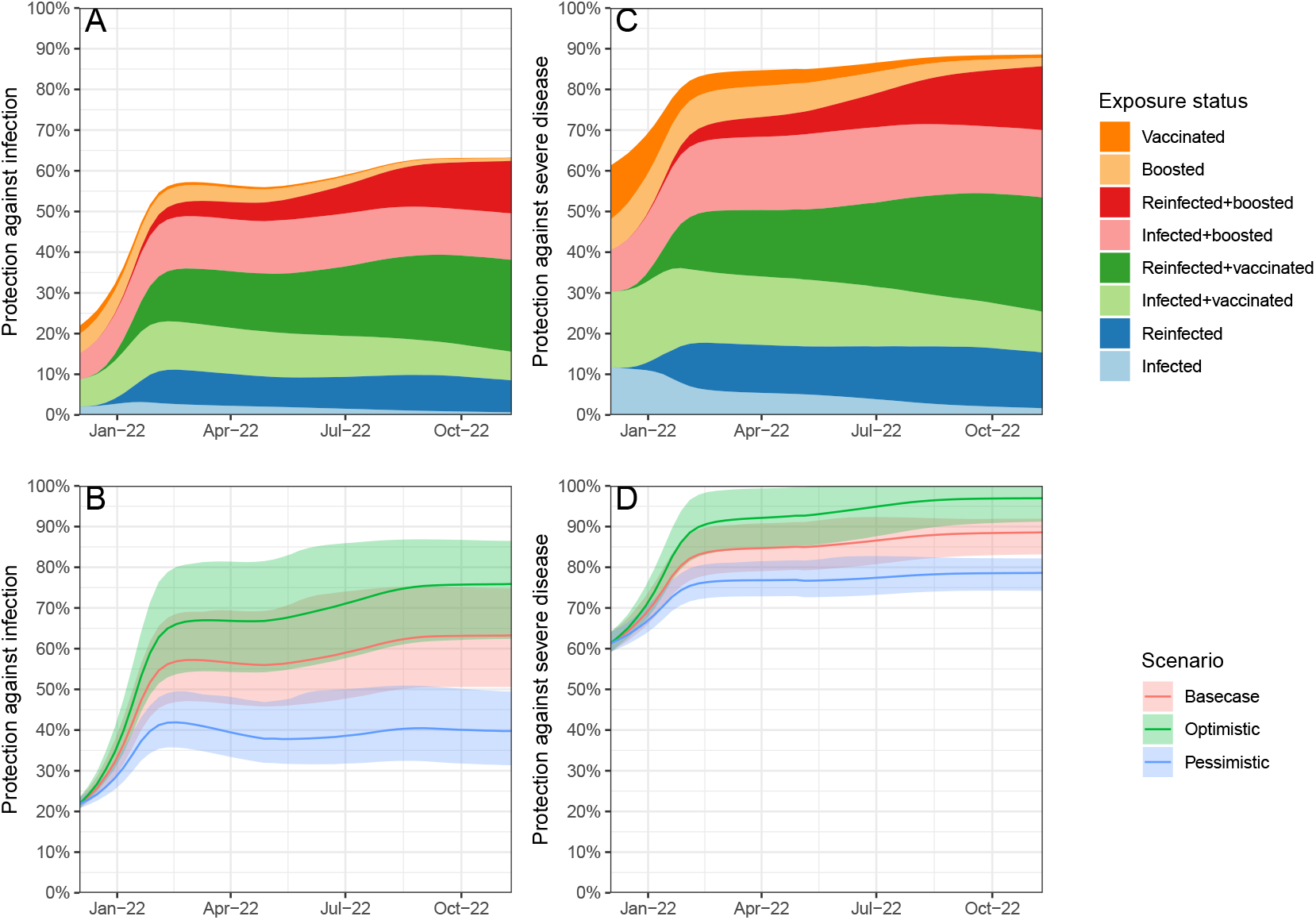
Population protection against infection and severe disease from the Omicron variant in the United States population between December 1, 2021 and November 9, 2022, by exposure state and waning scenario

At the state level, protection against infection ranged from 55% (Mississippi, 43%–72%) to 69% (Maine, 54%–83%) as of November 9, 2022. By this date, we estimated that half of the states had time trends in protection against infection that had started to decline, i.e., the existing immunity waned more quickly than new immunity was being accumulated (eFigure 3 in the Supplement).

Under the pessimistic waning scenario, national protection against infection on November 9, 2022 was 40% (31%–50%), while under the optimistic waning scenario, it was 76% (62%–87%) (Figure 3CD). Figure 4 shows state-level results for how population protection has increased from December 2, 2021, to March 3, 2022, and then to November 9, 2022. In all but eight states, population immunity against a new Omicron infection was estimated to have increased or stayed constant between the end of the December-February surge and the end of the study period. In more than one fourth of the counties the population immunity against a new Omicron infection decreased between March 3, 2022 and November 9, 2022 (eFigure 6).

**Figure 4:**
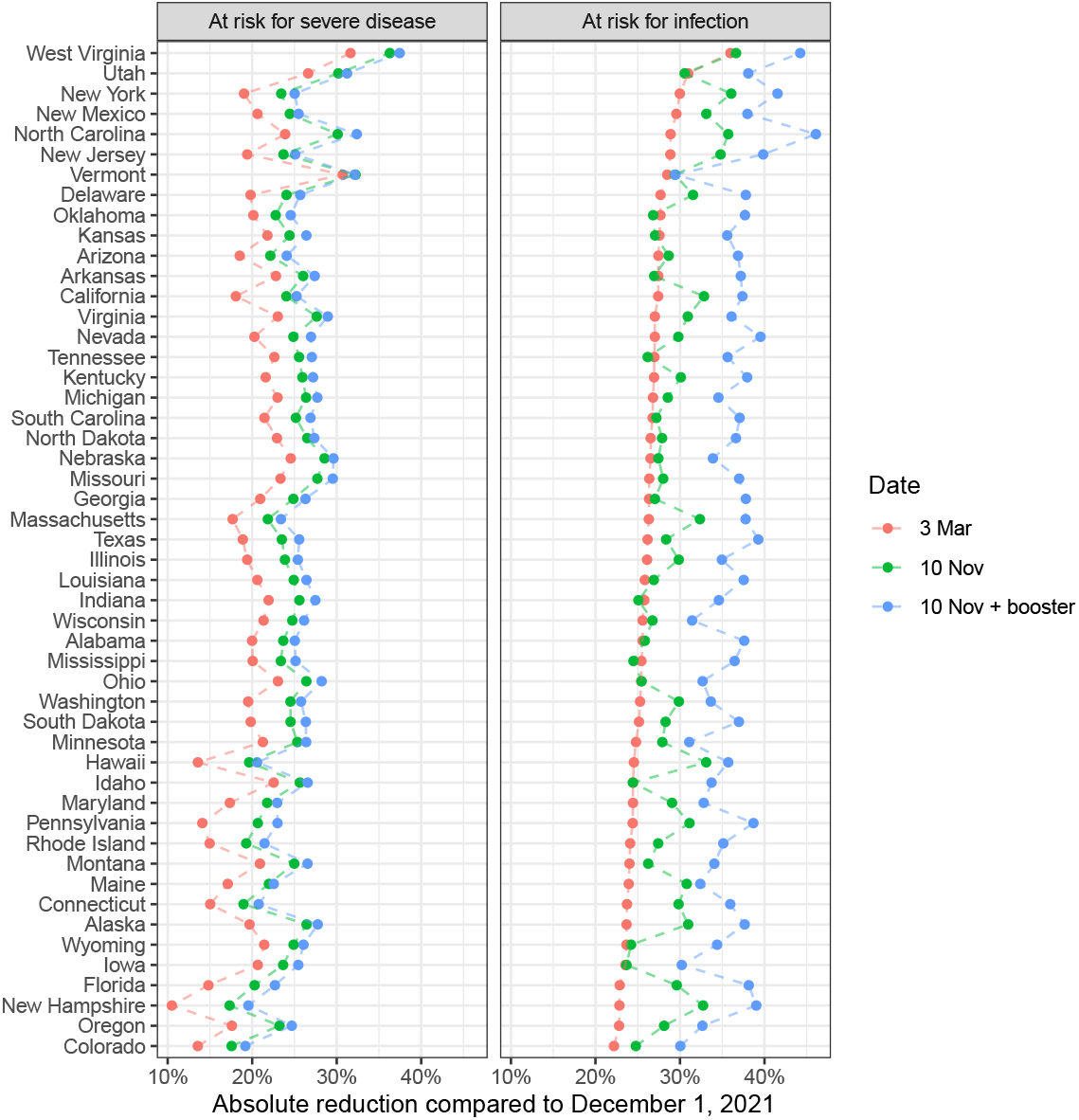
State-level changes in population protection against infection and severe disease from the Omicron variant between December 1, 2021 and November 9, 2022

### Population immunity against severe disease

Population protection against severe disease was estimated to have increased over the study period, both nationally (Figure 3CD) and within every state (Figure 4). Nationally, protection against severe disease resulting from a new Omicron infection rose from 61% (59%–64%) on December 2, 2021, to 84% (79%–90%) on March 3, 2022, to 89% (83%–91%) by November 9, 2022.

### Booster uptake scenarios

On November 9, 79% of the US population has initiated a vaccination series (range: 60% Wyoming – 95% Vermont and Maine), 34% has received a first booster (range: 20% North Carolina – 55% Vermont) and 11% has received a second booster (range: 5% North Carolina – 22% Vermont). We evaluated the impact on immunity if all states were able to increase coverage of first and second boosters to the levels seen in the best performing state (55% and 22% respectively, Vermont). We assumed initial vaccine coverage was saturated. In this hypothetical scenario, national protection against infection would increase 4 percentage points (state range, excluding Vermont: 1.6 (Maine) – 12 (Mississippi))), and protection against severe disease would go up by 1.1 percentage point (state range, excluding Vermont: 0.5 (Maine) – 2.4 (Florida)) (Figure 4).

## Discussion

In this study, we estimated time trends in effective protection against infection and severe disease caused by SARS-CoV-2 Omicron variants in the United States between December 2021 and November 2022. The results of this analysis show that effective protection against SARS-CoV-2 infection and severe disease on November 9, 2022 was substantially higher than on December 2, 2021, before the initial Omicron surge. We also found that population immunity in November 2022 was higher than in the immediate aftermath of the initial surge, with the impact of ongoing transmission and vaccination over the middle of 2022 outweighing the losses in protection due to waning of immunity, both nationally and for the large majority of US states.

Our estimate for national protection against a new Omicron infection (mean estimate 63% on November 9, 2022), is comparable to estimates produced by other studies (IHME:^24^ 58% against infection with BA.5 variant, 62% against infection with B.1.1 variant; and McKinsey:^25^ 75%). In addition to credible intervals for these estimates, we present a range of scenarios for the waning of immunity, rendering a wide range of potential levels of immunity. In particular, the pessimistic scenario presented shows that population protection against infection would be only around 40% nationally if prior Omicron infections do not offer additional protection against a new (sub)variant. This value is closer to the 22% population immunity estimated for the (at that time) new Omicron variants in December 2021^17^, as compared to population immunity values in November 2022.

By the end of our study period, over 94% of the US population was estimated to have been infected at least once with SARS-CoV-2, and a majority has been infected more than one time. Despite these high exposure numbers, there is still substantial population susceptibility to infection with an Omicron variant. In a hypothetical scenario of improved booster coverage, we found that increases in first and second booster uptake to the levels seen in the best-performing state would produce an appreciable improvement in population immunity, with greater relative impact for protection against infection vs. severe disease. This additional protection results from both the recovery of immunity lost due to waning and the increased effectiveness of the bivalent booster against Omicron infections^8^.

This study has several limitations. Firstly, vaccination data as reported by the CDC are known to be biased for some locations^26^, and there is no known alternative data source available that covers the full set of modelled locations accurately. Because of this bias, some modelled locations could have vaccination coverage that is higher than represented by reported data. While this could lead to underestimation of population immunity for these locations, this bias is likely small because, due to high infection rates, most of the population already has some level of immunity. Secondly, there is substantial uncertainty around the estimates of prior infectious exposure. Our analysis quantified many of the sources of uncertainty in the estimation approach, reflected in broad intervals for the estimated time course of infectious exposure in the analysis.

However, this analysis was based on an analytic model that makes structural assumptions about the natural history processes of COVID-19 and how these change across states and counties (described in detail in Chitwood, Russi et al 2021^12^), not all of which can be represented in the uncertainty analysis. Thirdly, we did not distinguish the levels of immune evasion achieved by successive Omicron-subvariants. Similarly, we did not distinguish the level of immunity conferred by different pre-Omicron variants, or different vaccine products. In the assignment of exposure states, we only explicitly make assumptions about the joint probability of vaccination and first infection, and do not account for behavior the changes the probability of changing exposure state (e.g., not getting boosted right after an infection).

A substantial fraction of the US population was infected with SARS-CoV-2 infection during December 2021 – February 2022, increasing population immunity against Omicron variants in the U.S. population. This level of immunity continued to increase slowly over the course of 2022, as newly-acquired immunity from ongoing transmission and vaccination outweighed the effects immune waning for the majority of modelled locations. Despite the high level of protection at the beginning of the 2022-2023 winter, risk of reinfection and subsequent severe disease remains present. A more transmissible or immune evading (sub)variant, changes in behavior, or ongoing waning of immunity could lead to a new SARS-CoV-2 wave, as was observed at the end of 2021.

## Data Availability

All data used in the main analysis are available from The Covid Tracking Project, Johns Hopkins CSSE, CDC, Healthdata.gov, US Census Bureau, Ipsos and a JAMA publication.

## Author contributions

TC, NAM and JAS conceived and supervised the project. TC, NAM and JAS acquired funding. FK wrote the model code, drafted the original manuscript and visualized the results. FK and MR curated the data and executed the analysis. All authors contributed to the development of the methodology, and reviewed and edited the original manuscript.

