## Supplement for "Changes in population immunity against infection and severe disease from SARS-CoV-2 Omicron variants in the United States between December 2021 and November 2022"

**Contents:**

eMethods

eFigure 1: Immunological exposure in the United States population between December 1, 2021 and November 9 2022, by exposure state

eFigure 2: Immunological exposure for each state in the United States between December 1, 2021 and November 9 2022, by exposure state

eFigure 3: Population protection against infection with the Omicron variant in each of the United States population between December 1, 2021 and November 9 2022, by waning scenario

eFigure 4: Population protection against severe disease from the Omicron variant in each of the United States population between December 1, 2021 and November 9 2022, waning scenario

eFigure 5: Population protection against infection with the Omicron variant on November 9, 2022 in each US county

eFigure 6: Increase in population protection against infection with the Omicron variant between March 3, 2022 and November 9 2022 in each US county

eTable 1: Exposure states and definitions

eTable 2: Levels of initial protection and rates of waning of protection against infection and severe disease by exposure state

**eMethods**

**Adaptations to the published model for infection estimates**

The analyses in this manuscript use estimates of infection that are generated using a published model that synthesizes reported case and death data[1]. This model is used for daily nowcasting of infections and has been updated in July 2022 to account for the Omicron variants. These changes are described in the changelog on the webpage www.covidestim.org and are repeated here for completeness:

- Hospitalizations: With the Omicron variant, the IFR has decreased relative to previous variants (fewer deaths per 100K infections). This caused problems for fitting the model – with relatively few deaths from many modelled locations, it became difficult to produce precise estimates of COVID-19 burden. The revised model is estimated using data on COVID-19 hospitalizations, instead of COVID-19 deaths. The hospitalizations data are extracted from healthdata.gov[2]. This data set contains weekly data on hospital admissions for each facility in the US. We aggregate these data, first the the Health Service Area level, then to the county level and finally to the state level (detailed description here). 'Confirmed Hospital Admissions (Admissions Confirmed Adults)' is the variable of interest for the covidestim model. This variable is censored for observations of 1-3 hospitalizations per facility. We use the lower bound of the aggregate variables. *i.e.,* if a facility reports censored data in a week, we assume 1 new admission has occurred in that facility.
- Weekly time-step: The hospitalizations data are reported on a weekly basis. Therefore, we have adapted the covidestim model to use a weekly timestep. This change also reduces the computational time required to fit the model. The daily case data are summed to match the weeks of the hospitalizations. All time dependent prior distributions and delay distributions in the model have been converted to match the weekly timescale by multiplying the scale parameter by 7.
- First date of model: the estimation in this update starts on December 1, 2021, to align with the beginning of the Omicron wave. This allows the model to estimate transition probabilities specific to the variant distribution after December 1, 2021.
- Reinfection. New in the covidestim model is the possibility of reinfection. In the previous version of the model, the new infections on each day were subtracted from the susceptible population; implying that an individual could not be reinfected. Currently, three measures of protection are incorporated in the model.
    - Immunity from infection: In this new implementation, we do not assume that an infection prevents reinfection. Rather, we assume that following an infection, immunity wanes progressively as described below ('Waning of immunity').

- Historic immunity: each covidestim run is initialized with an estimate of the population protection against infection with the Omicron variant on December 1, 2021. This estimate is obtained from the calculations from Klaassen et al. 2022[3].
- Booster immunity: the data for any boosters and first vaccinations administered after December 1, 2021, are extracted from the CDC data dashboard for counties and for states. For states where the cumulative boosters or first vaccinations data exceeds the population size (New Hampshire and Rhode Island in the latest inspection), we impute the average of the booster/first vaccinations data from neighboring states. If there are no neighbors, the average of all valid states is used. For counties where the cumulative booster or first vaccination data exceeds the population size, or where the booster data is missing, the state average is imputed. We assume the booster starts at an efficacy of 80% and then wanes exponentially.

- Waning of immunity. All three measures of immunity, have the same associated exponential waning curve associated with them with a median of 120 days (half of the people with an infection will be eligible for reinfection after 120 days).

In addition to the above changes, one additional change was made to the model. We assumed that the probability to progress to severe disease for those individuals with a repeated infection (not first infection). To reflect this assumption, we reduced the total infection fatality ratio for those reinfected by 90%. That is, a constant protection of 90% against death was assumed.

**Estimation of infections**

Estimates of immunological exposure and immunity from Klaassen et al. (2022) were used to initialize the covidestim model. These estimates were obtained using different data than used in the current manuscript. First, we now use the CDC vaccination data[4], because this is, to our knowledge, the only dataset that reports the number of second booster doses in addition to the primary vaccination series and the first booster. Second, in Klaassen et al, the under 12 population was assumed to not have access to vaccinations, and the vaccination data was adjusted to reflect this assumption. In the current manuscript, we do not make this assumption. As a consequence, the number of vaccinated individuals in each location is not smoothly transitioning from the initialization point on December 1, 2021, to the currently used timeseries. We corrected for these discrepancies as follows:

- If the cumulative vaccinations in the initializations data was LOWER than the observed cumulative vaccination data on December 1, we included the difference in the number of vaccinations on December 1.
- If the cumulative vaccinations in the initializations data was HIGHER than the observed cumulative vaccination data on December 1, we set the new vaccinations to 0, until the observed cumulative vaccinations exceeded the initialized data.

**eTable 1. Definitions of exposures states**

| Exposure state | Description |
|---|---|
| Naïve | Never infected or vaccinated |
| Infected (pre-omicron) | Infected before December 1, 2021. That is, assumed to be infected with a pre-omicron variant. |
| Infected (omicron) | Infected after December 1, 2021. That is, assumed to be infected with the Omicron variant. |
| Reinfected | Reinfections always have occurred only after December 1, 2021, and are therefore assumed to be infections with the Omicron variant. |
| Infected (pre-omicron) and vaccinated | Infected before December 1, 2021 (with a pre-Omicron variant), and initiated a primary vaccination series. |
| Infected (omicron) and vaccinated | Infected after December 1, 2021 (with the Omicron variant), and initiated a primary vaccination series. |
| Reinfected (pre-omicron) and vaccinated | Infected after December 1, 2021 (with the Omicron variant), and initiated a primary vaccination series. |
| Infected (pre-omicron) and boosted | Infected before December 1, 2021 (with a pre-Omicron variant), completed a primary vaccination series and received one additional booster dose. |
| Infected (omicron) and boosted | Infected after December 1, 2021 (with the Omicron variant), completed a primary vaccination series and received one additional booster dose. |
| Reinfected (pre-omicron) and boosted | Infected after December 1, 2021 (with the Omicron variant), completed a primary vaccination series and received one additional booster dose. |
| Infected (pre-omicron) and double boosted | Infected before December 1, 2021 (with a pre-Omicron variant), completed a primary vaccination series and received two additional booster doses. |
| Infected (omicron) and double boosted | Infected before December 1, 2021 (with the Omicron variant), completed a primary vaccination series and received two additional booster doses. |
| Reinfected (pre-omicron) and double boosted | Infected before December 1, 2021 (with the Omicron variant), completed a primary vaccination series and received two additional booster doses. |
| Double boosted | Completed a primary vaccination series and received two additional booster doses. |
| Boosted | Completed a primary vaccination series and received one additional booster dose. |
| Vaccinated | Initiated a primary vaccination series. |

**eTable 2. Levels of initial protection and rates of waning of protection against infection and severe disease by exposure state**

Footnote: The initial level of immunity afforded by a given exposure was assumed to be ordered as follows, from low to high immunity: vaccination (first dose), booster, pre-Omicron infection, pre-Omicron infection + vaccination, pre-Omicron infection + booster, Omicron (re)infections, Omicron (re)infections + vaccination, Omicron (re)infections + booster. For the protection against severe disease, we did not differentiate the initial protection afforded by Omicron and pre-Omicron infections, as evidence suggests the protection against severe disease is consistent

| Exposure state | Initial protection against infection | Monthly reduction in protection against infection | Initial protection against severe disease | Monthly reduction in protection against severe disease |
|---|---|---|---|---|
| Infected (pre-Omicron) | 60% | 8% | 90% | 0.8% |
| Infected (Omicron) / reinfected | 70% | 8% | 90% | 0.8% |
| Infected (pre-omicron) and vaccinated | 70% | 4% | 90% | 0% |
| Infected / reinfected (omicron) and vaccinated | 80% | 2% | 90% | 0% |
| Infected (pre-omicron) and boosted / double boosted | 75% | 4% | 95% | 0% |
| Infected / reinfected (omicron) and boosted / double boosted | 85% | 2% | 95% | 0% |
| Boosted / double boosted | 50% | 8% | 85% | 0.8% |
| Vaccinated | 30% | 8% | 80% | 0.8% |

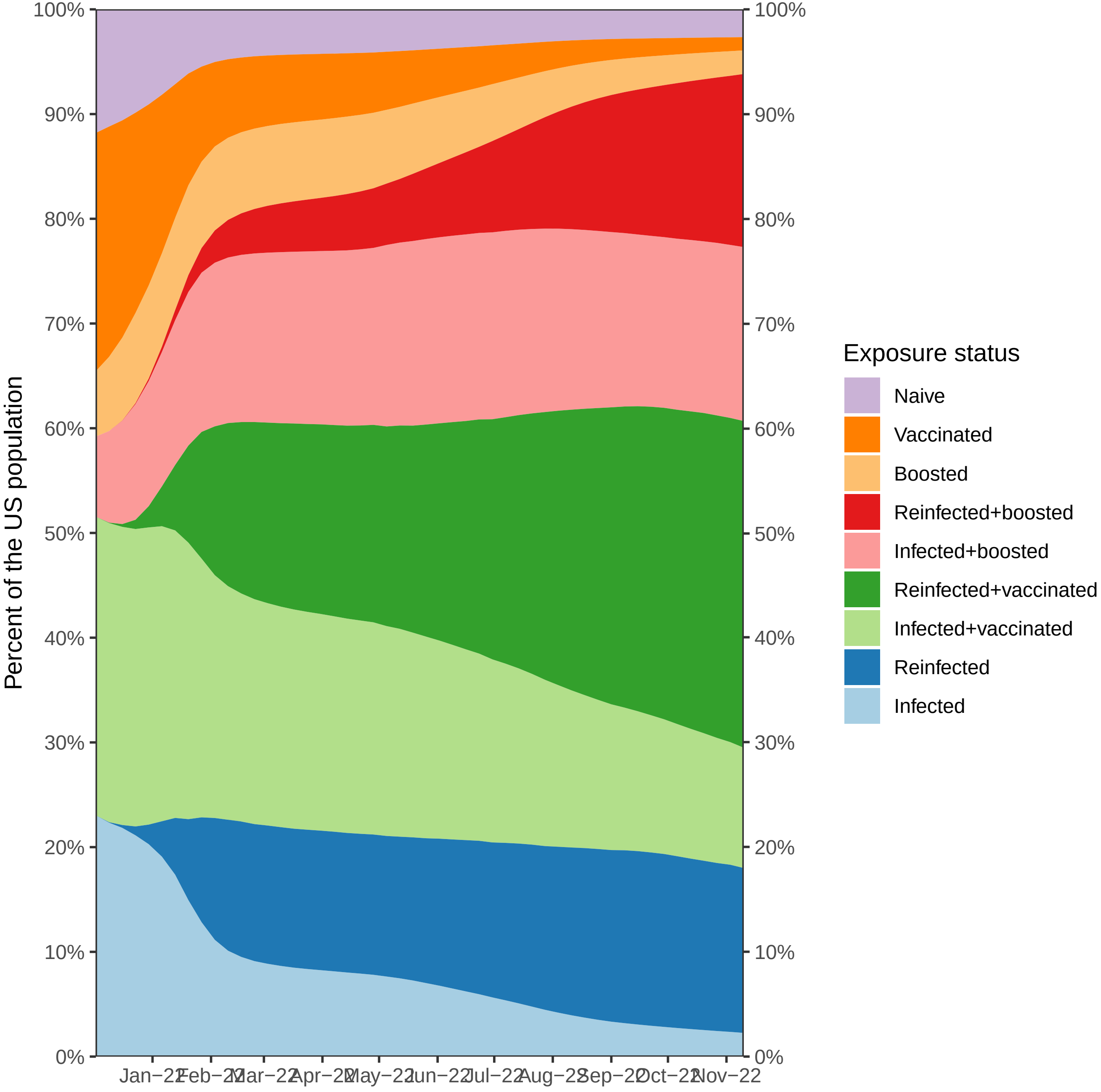

Percent of the US population
100%
90%
80%
70%
60%
50%
40%
30%
20%
10%
0%
Jan−22
Feb−22
Mar−22
Apr−22
May−22
Jun−22
Jul−22
Aug−22
Sep−22
Oct−22
Nov−22
Exposure status
Naive
Vaccinated
Boosted
Reinfected+boosted
Infected+boosted
Reinfected+vaccinated
Infected+vaccinated
Reinfected
Infected

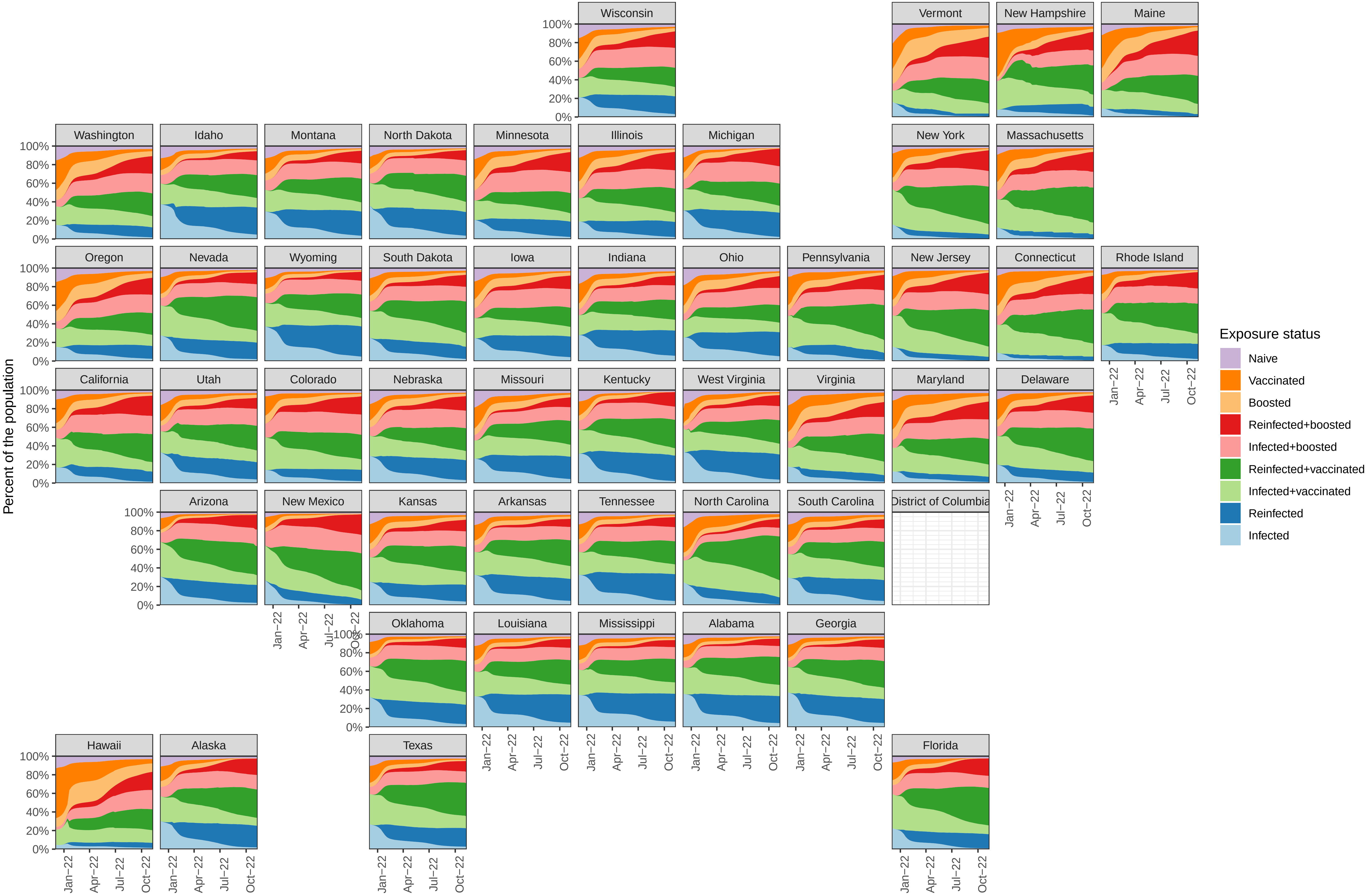
Wisconsin
Vermont
New Hampshire
Maine
Washington
Idaho
Montana
North Dakota
Minnesota
Illinois
Michigan
New York
Massachusetts
Oregon
Nevada
Wyoming
South Dakota
Iowa
Indiana
Ohio
Pennsylvania
New Jersey
Connecticut
Rhode Island
California
Utah
Colorado
Nebraska
Missouri
Kentucky
West Virginia
Virginia
Maryland
Delaware
Arizona
New Mexico
Kansas
Arkansas
Tennessee
North Carolina
South Carolina
District of Columbia
Oklahoma
Louisiana
Mississippi
Alabama
Georgia
Hawaii
Alaska
Texas
Florida
100%
80%
60%
40%
20%
0%
Jan−22
Apr−22
Jul−22
Oct−22
Percent of the population
Exposure status
Naive
Vaccinated
Boosted
Reinfected+boosted
Infected+boosted
Reinfected+vaccinated
Infected+vaccinated
Reinfected
Infected

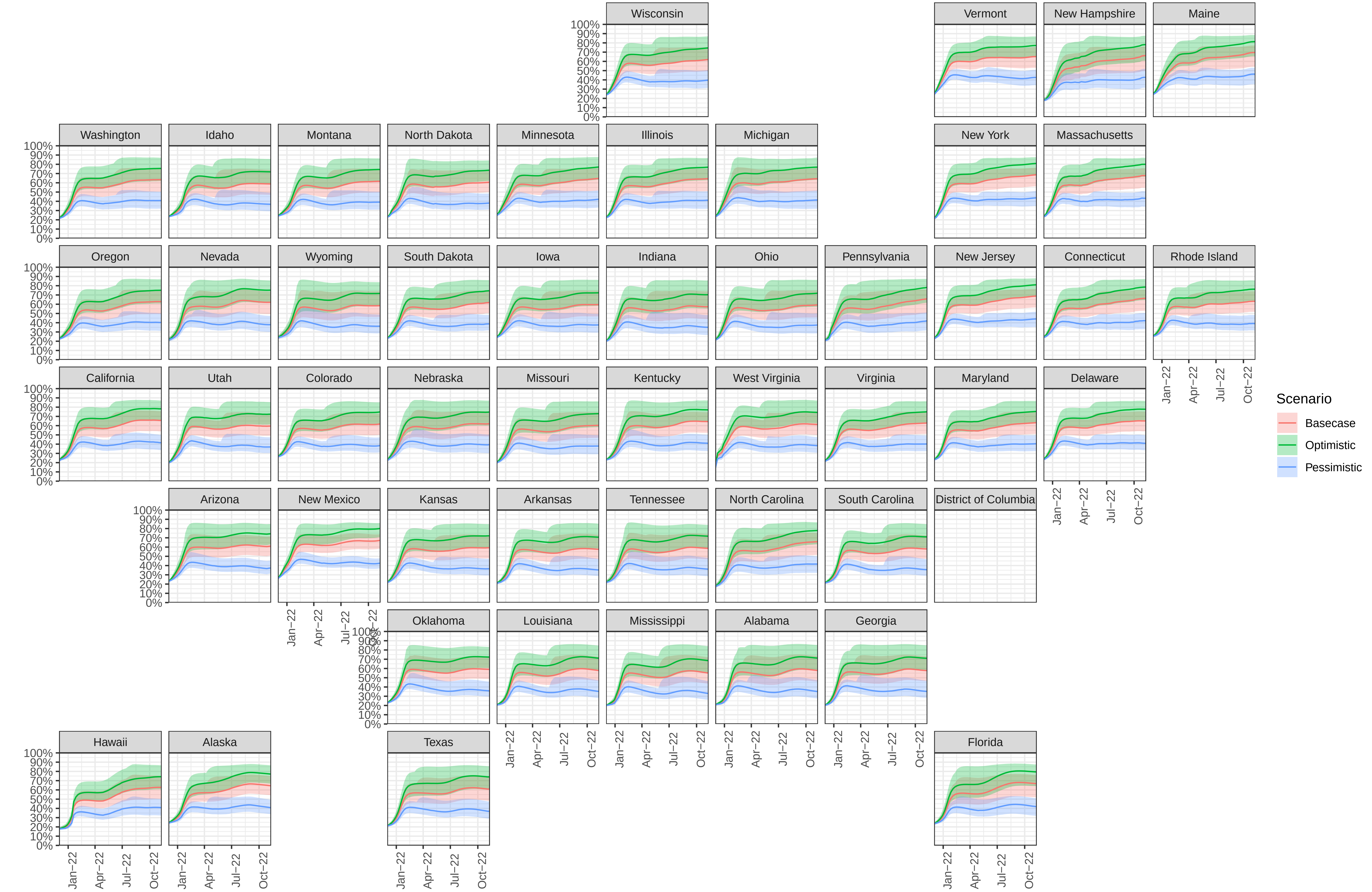
Wisconsin
Vermont
New Hampshire
Maine
Washington
Idaho
Montana
North Dakota
Minnesota
Illinois
Michigan
New York
Massachusetts
Oregon
Nevada
Wyoming
South Dakota
Iowa
Indiana
Ohio
Pennsylvania
New Jersey
Connecticut
Rhode Island
California
Utah
Colorado
Nebraska
Missouri
Kentucky
West Virginia
Virginia
Maryland
Delaware
Arizona
New Mexico
Kansas
Arkansas
Tennessee
North Carolina
South Carolina
District of Columbia
Oklahoma
Louisiana
Mississippi
Alabama
Georgia
Hawaii
Alaska
Texas
Florida
100%
90%
80%
70%
60%
50%
40%
30%
20%
10%
0%
Jan−22
Apr−22
Jul−22
Oct−22
Scenario
Basecase
Optimistic
Pessimistic

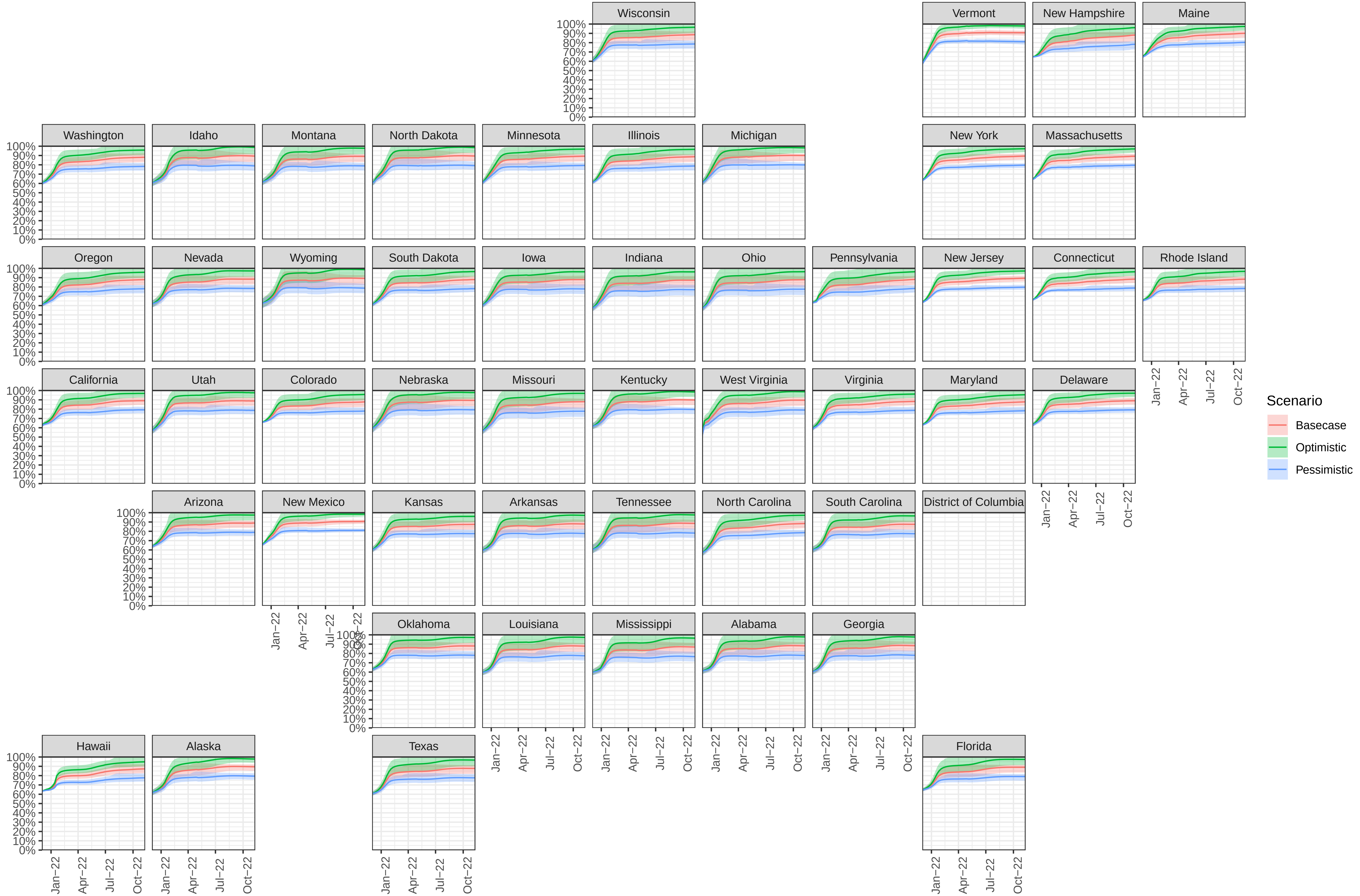

Wisconsin
Vermont
New Hampshire
Maine
Washington
Idaho
Montana
North Dakota
Minnesota
Illinois
Michigan
New York
Massachusetts
Oregon
Nevada
Wyoming
South Dakota
Iowa
Indiana
Ohio
Pennsylvania
New Jersey
Connecticut
Rhode Island
California
Utah
Colorado
Nebraska
Missouri
Kentucky
West Virginia
Virginia
Maryland
Delaware
Arizona
New Mexico
Kansas
Arkansas
Tennessee
North Carolina
South Carolina
District of Columbia
Oklahoma
Louisiana
Mississippi
Alabama
Georgia
Hawaii
Alaska
Texas
Florida
100%
90%
80%
70%
60%
50%
40%
30%
20%
10%
0%
Jan−22
Apr−22
Jul−22
Oct−22
Scenario
Basecase
Optimistic
Pessimistic

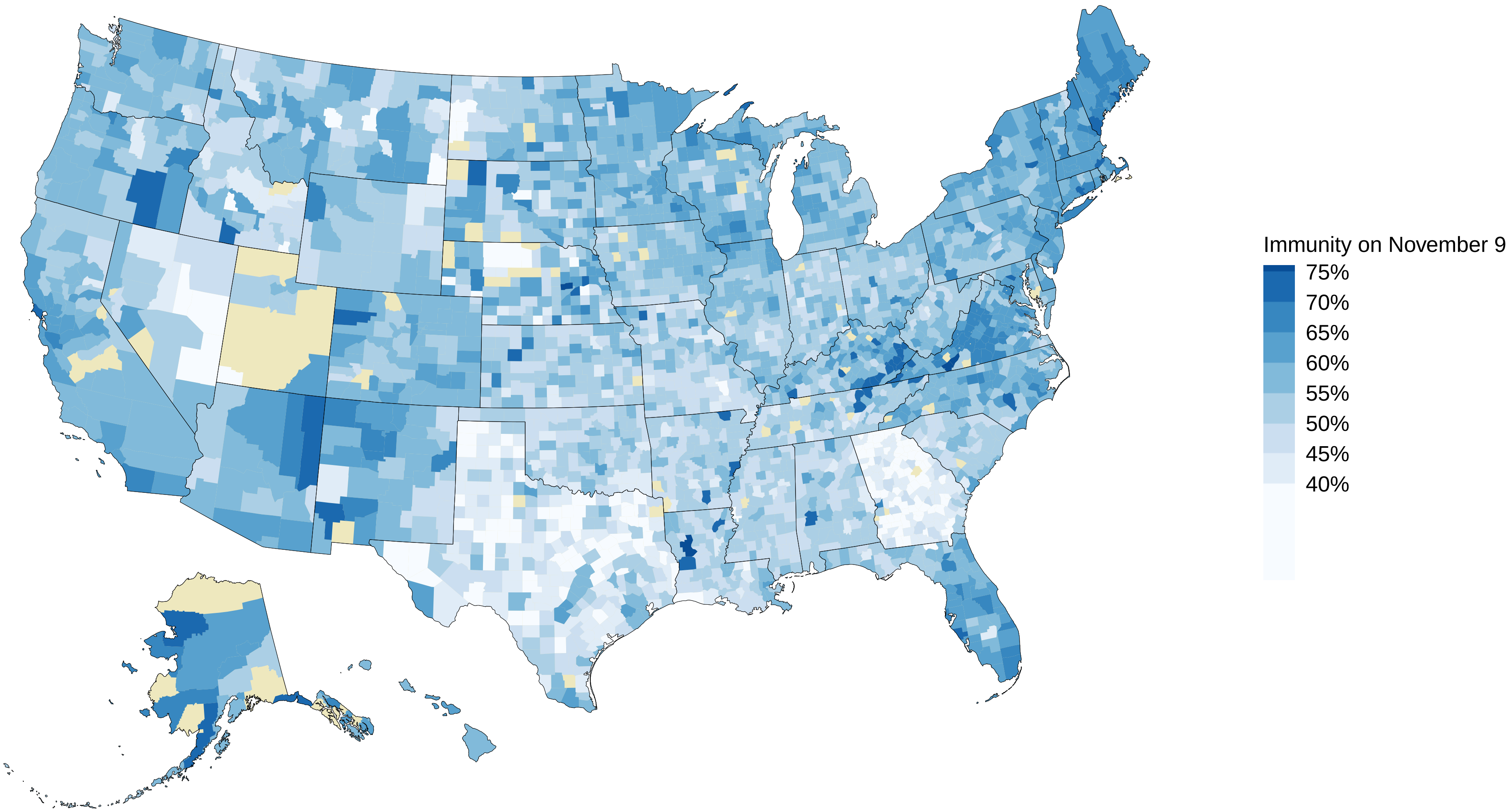
Immunity on November 9
75%
70%
65%
60%
55%
50%
45%
40%

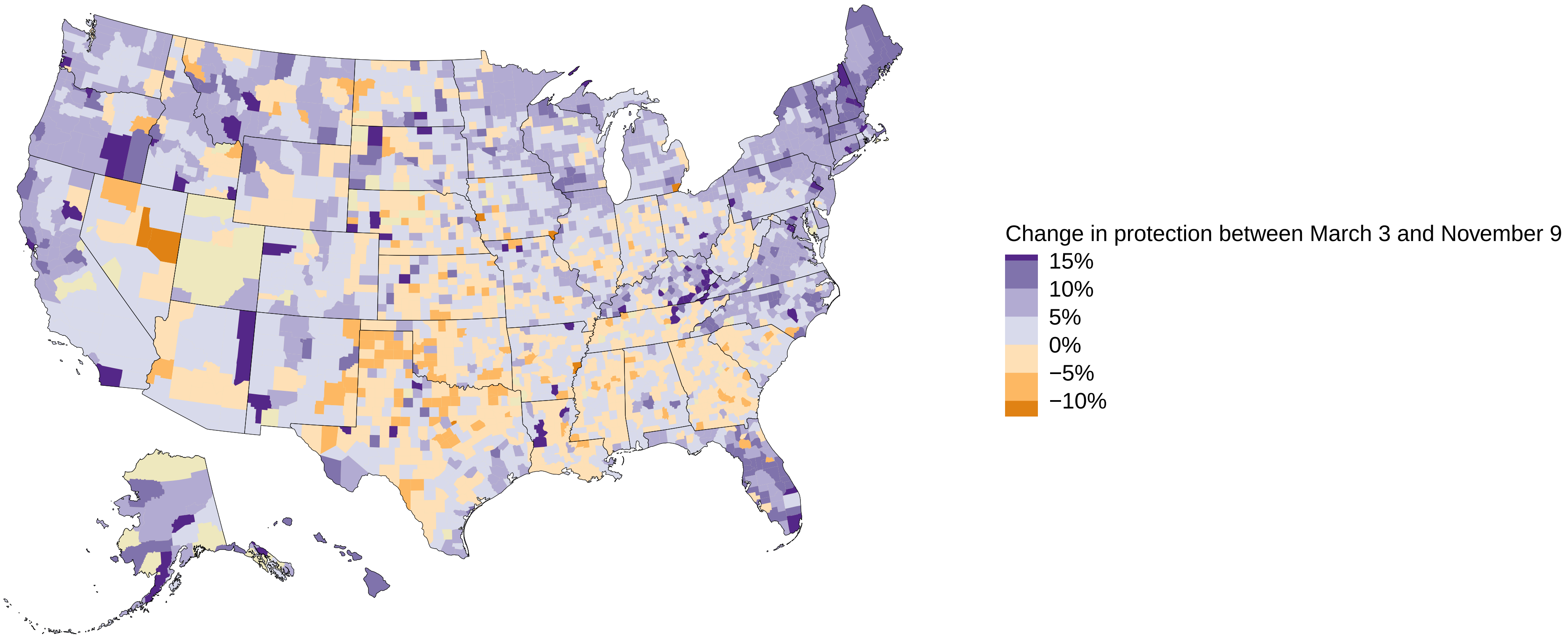
Change in protection between March 3 and November 9
15%
10%
5%
0%
−5%
−10%
